# Value of radiomics features from adrenal gland and periadrenal fat CT images predicting COVID-19 progression

**DOI:** 10.1101/2021.01.03.21249183

**Authors:** Mudan Zhang, Xuntao Yin, Wuchao Li, Yan Zha, Xianchun Zeng, Xiaoyong Zhang, Jingjing Cui, Jie Tian, Rongpin Wang, Chen Liu

**Affiliations:** Guizhou University School Of Medicine, Guiyang, Guizhou province, 550000, China; Department of Radiology, Guizhou Provincial People’s Hospital, Guiyang, Guizhou province, 550002, China; Guizhou Provincial Key Laboratory of Intelligent Medical Image Analysis and Precision Diagnosis, Guizhou Provincial People’s Hospital, Guiyang, Guizhou province, 550002, China; Department of Nephrology, Guizhou Provincial People’s Hospital, Guiyang, Guizhou province, 550002, China; NHC Key Laboratory of Pulmonary Immunological Diseases, Guizhou University People’s Hospital, Guiyang, Guizhou province, 550002, China; Shanghai United Imaging Intelligence, Co., Ltd., Shanghai, 201807, China; Key Laboratory of Molecular Imaging, Chinese Academy of Sciences, Beijing, 100190, China; Department of Radiology, Southwest Hospital, Third Military Medical University(Army Medical University), Chongqing, 400038, China

**Keywords:** adrenal gland, periadrenal fat, auto-segmentation, COVID-19, radiomics

## Abstract

**Background:** Value of radiomics features from the adrenal gland and periadrenal fat CT images for predicting disease progression in patients with COVID-19 has not been studied.

**Methods:** A total of 1,245 patients (685 moderate and 560 severe patients) were enrolled in a retrospective study. We proposed 3D V-Net to segment adrenal glands in onset CT images automatically, and periadrenal fat was obtained using inflation operation around the adrenal gland. Next, we built a clinical model (CM), three radiomics models (adrenal gland model [AM], periadrenal fat model [PM], and fusion of adrenal gland and periadrenal fat model [FM]), and radiomics nomogram (RN) after radiomics features extracted to predict disease progression in patients with COVID-19.

**Results:** The auto-segmentation framework yielded a dice value of 0.79 in the training set. CM, AM, PM, FM, and RN obtained AUCs of 0.712, 0.692, 0.763, 0.791, and 0.806, respectively in the training set. FM and RN had better predictive efficacy than CM (*P* < 0.0001) in the training set. RN showed that there was no significant difference in the validation set (mean absolute error [MAE] = 0.04) and test set (MAE = 0.075) between predictive and actual results. Decision curve analysis showed that if the threshold probability was more than 0.3 in the validation set or between 0.4 and 0.8 in the test set, it could gain more net benefits using RN than FM and CM.

**Conclusion:** Radiomics features extracted from the adrenal gland and periadrenal fat CT images may predict progression in patients with COVID-19.

**Funding:** This study was funded by Science and Technology Foundation of Guizhou Province (QKHZC [2020]4Y002, QKHPTRC [2019]5803), the Guiyang Science and Technology Project (ZKXM [2020]4), Guizhou Science and Technology Department Key Lab. Project (QKF [2017]25), Beijing Medical and Health Foundation (YWJKJJHKYJJ-B20261CS) and the special fund for basic Research Operating Expenses of public welfare research institutes at the central level from Chinese Academy of Medical Sciences (2019PT320003).

## Introduction

Coronavirus disease 2019 (COVID-19) has caused serious public health problems, which have been much worse than those caused by SARS in 2002–2003. Until December 1, 2020, WHO reported 62,363,527 confirmed patients and 1,456,687 deaths globally (https://covid19.who.int/). Although a diagnosis and treatment protocol suitable for the national situation has been enacted in each country, the consensus is to separate moderate patients from severe patients who need special care. We must predict early disease progression and accurately identify which patients are at risk of developing a severe or critical disease. This is not only conducive to the early disease control but will also effectively allocate medical resources globally.

Because it was unclear that the polymerase chain reaction (PCR) technique(1), clinical symptoms, and laboratory tests were correlated with progression in patients with COVID-19 (2), researchers started exploring predicting the disease prognosis of COVID-19 combining artificial intelligence (AI) technology and CT images. Zhang et al. (3) constructed a comprehensive system for accurate diagnosis, quantitative measurements, and prognoses of COVID-19 pneumonia using chest CT images and AI technology. They reported more than 0.95 of AUC, both in training and validation sets. The results proved that the combination of chest CT and AI technology is a potential tool to predict disease progression in patients with COVID-19 using chest CT images and clinical indicators based on AI technology. Most studies have focused on the relationship between features from chest CT images with disease progression and ignored the endocrine system that played an important role in disease progression (4). The endocrine system was largely unnoticed during the outbreak of SARS. It has been proved that damage and changes in endocrine functions were important predictors of morbidity and mortality during the previous SARS outbreak (5). The endocrine system was largely unnoticed during the outbreak of SARS. It has been proved that damage and changes in endocrine functions were important predictors of morbidity and mortality during the previous SARS outbreak (6). However, studies showing whether the prognosis of patients with COVID-19 can be affected by direct damage or indirectly change of adrenal glands or periadrenal fat are lacking.

The adrenal gland is a key component of the body’s stress and immune system that is composed of the hypothalamic-pituitary-adrenal axis and the sympathetic-adrenal-medullary system (7). The adrenal gland integrates the cortex, which secretes glucocorticoids plus catecholamines, and the medulla, which secretes catecholamines within one organ capsule. The pituitary gland mostly regulates adrenal cortex function through adrenocorticotropic hormone (ACTH) and aldosterone, an important part of the renin-angiotensin system. The adrenal medulla also secretes catecholamines in response to the sympathetic nervous system (SNS). Entire system and the organs mentioned above are closely related to the immune-inflammatory state of COVID-19 patients (8). Moreover, the adrenal gland can be changed not only by receiving a direct attack from SARS-CoV-2 through angiotensin-converting enzyme-2 (ACE2) (9) but also by adapting to physiological needs and pathological conditions because it has an astonishing regenerative capacity to adjust to unique micro-environments based on an extensive vascular network (10). Therefore, we hypothesized that adrenal gland changes could occur due to its adaptive mechanism and SARS-CoV-2 invasion. The periadrenal fat was bound to be affected as the closest tissue around the adrenal gland. Fundamental changes can be detected by extracting radiomics features from chest CT images using AI technology. Radiomics is an emerging technique that can convert images difficult for human eyes to distinguish into high-throughput quantitative features that may reflect potential pathological and physiological states (11). Manual segmentation is the first and key step of successfully constructing a radiomics model, but it is time-consuming and error-prone because of repeated labor. Combining the auto-segmentation framework based on AI technology with manual revision improves both efficiency and quality (12). Cascaded V-Nets have been used to segment brain tumors with good results (13), which could be further improved if incorporated with extra information, such as coarse localization.

We therefore constructed an auto-segmentation framework and developed a radiomics nomogram (RN) to predict progression in patients with COVID-19 by integrating radiomics features from adrenal glands and periadrenal fat CT images with clinical indicators and to determine that radiomics features extracted from the adrenal gland and periadrenal fat CT images were related to progression in patients with COVID-19.

## Results

### Patient characteristics

A total of 1,245 patients (685 moderate and 560 severe patients) were enrolled in the retrospective study from two hospitals (1,209 patients from one and 36 patients from the other). The data of 1,209 patients were formed as our training and validation sets, including 672 patients who suffered from moderate COVID-19 and 537 who suffered from severe COVID-19. The patient characteristics in training and validation sets are listed in Table 1. No significant differences were observed between the training and validation set in sex (*P* = 0.238). lnterleukin 6 (IL-6) levels, white blood cell count (WBC), lymphocyte count (L), neutrophil count (N), C-reactive protein (CRP), procalcitonin (PCT), prothrombin time (PT), D dimer (DD), blood sugar (BS), glutamic oxaloacetic transaminase (AST), albumin (ALB), direct bilirubin (D-Bil), B-type natriuretic peptide (BNP), lactate dehydrogenase (LDH), and creatine kinase-MB (CK-MB) differed significantly between moderate and severe pneumonia sets both in training and validation sets (*P* < 0.05).

**Table 1.**
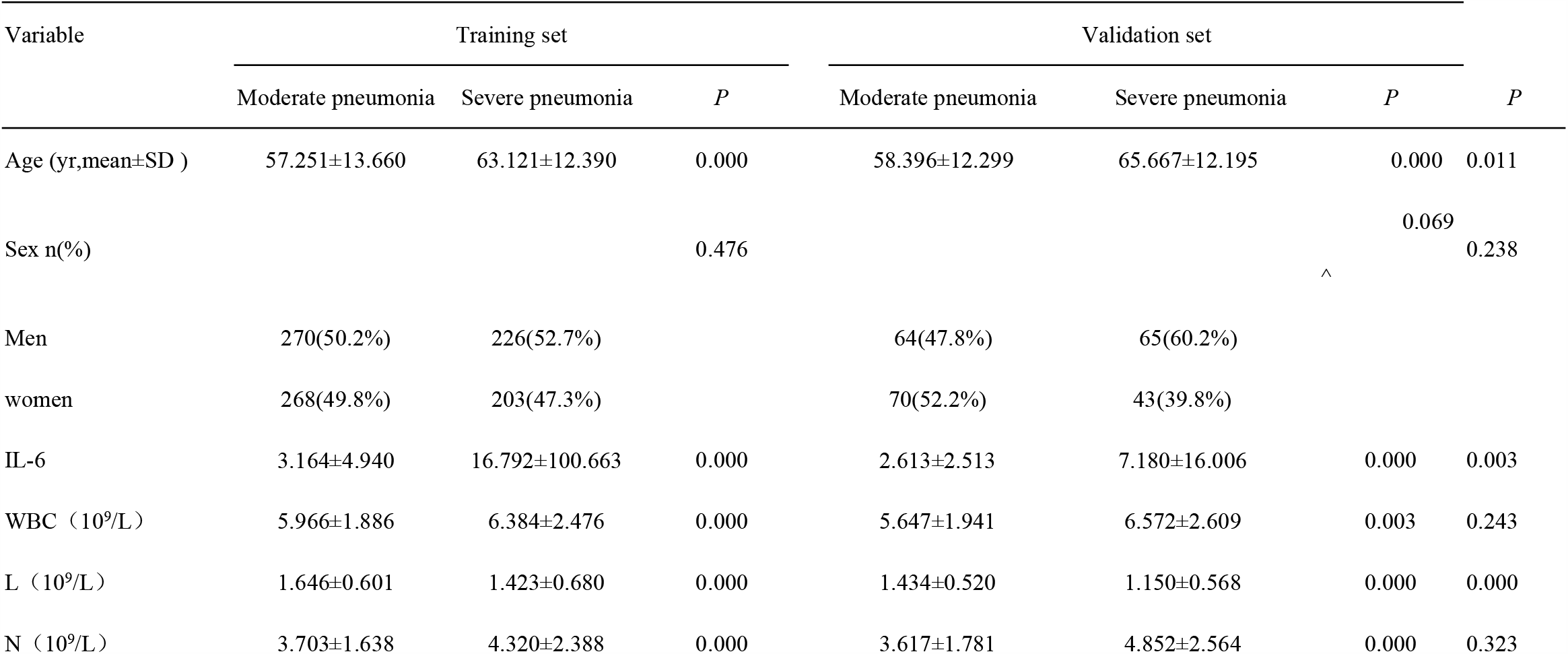
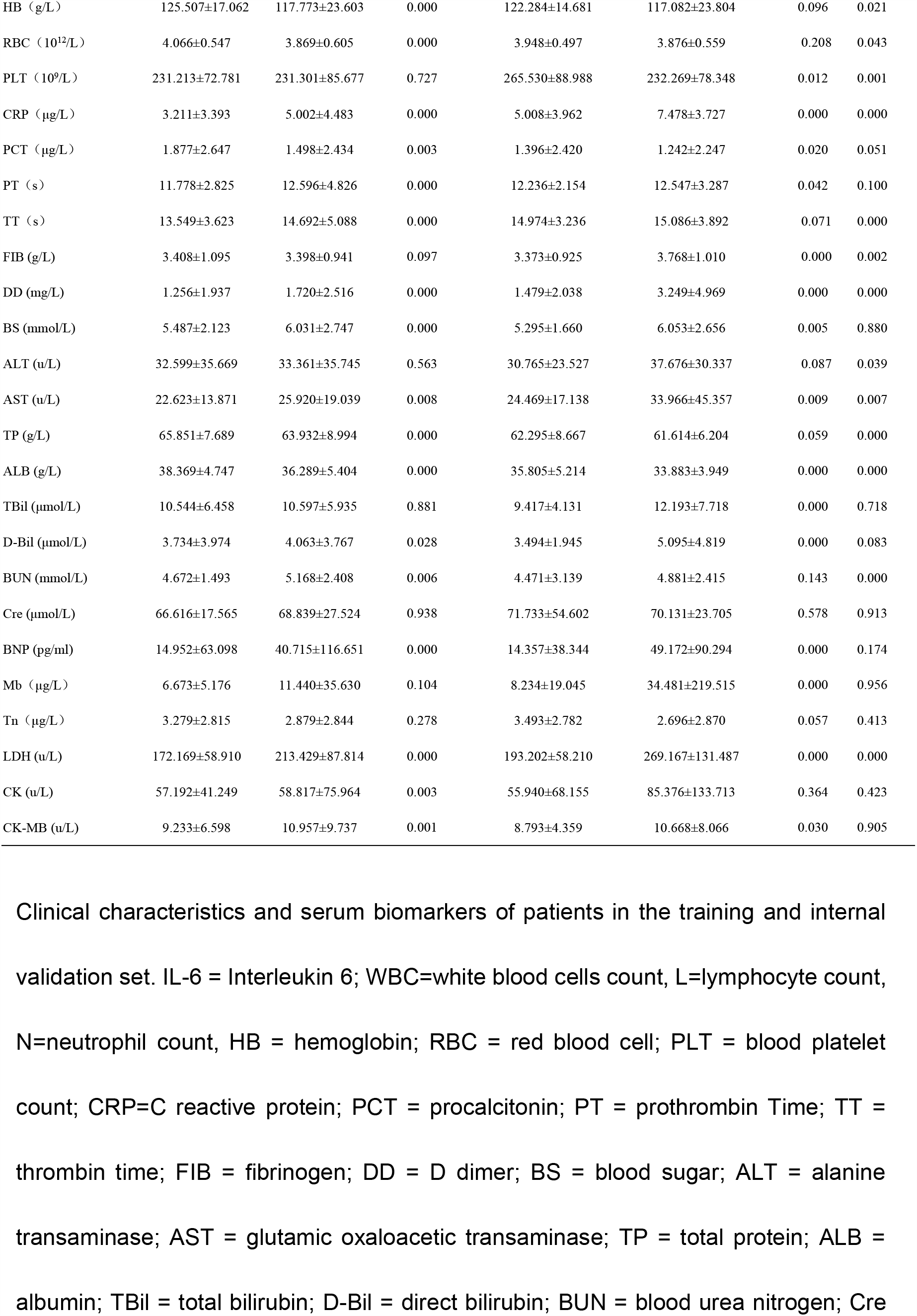
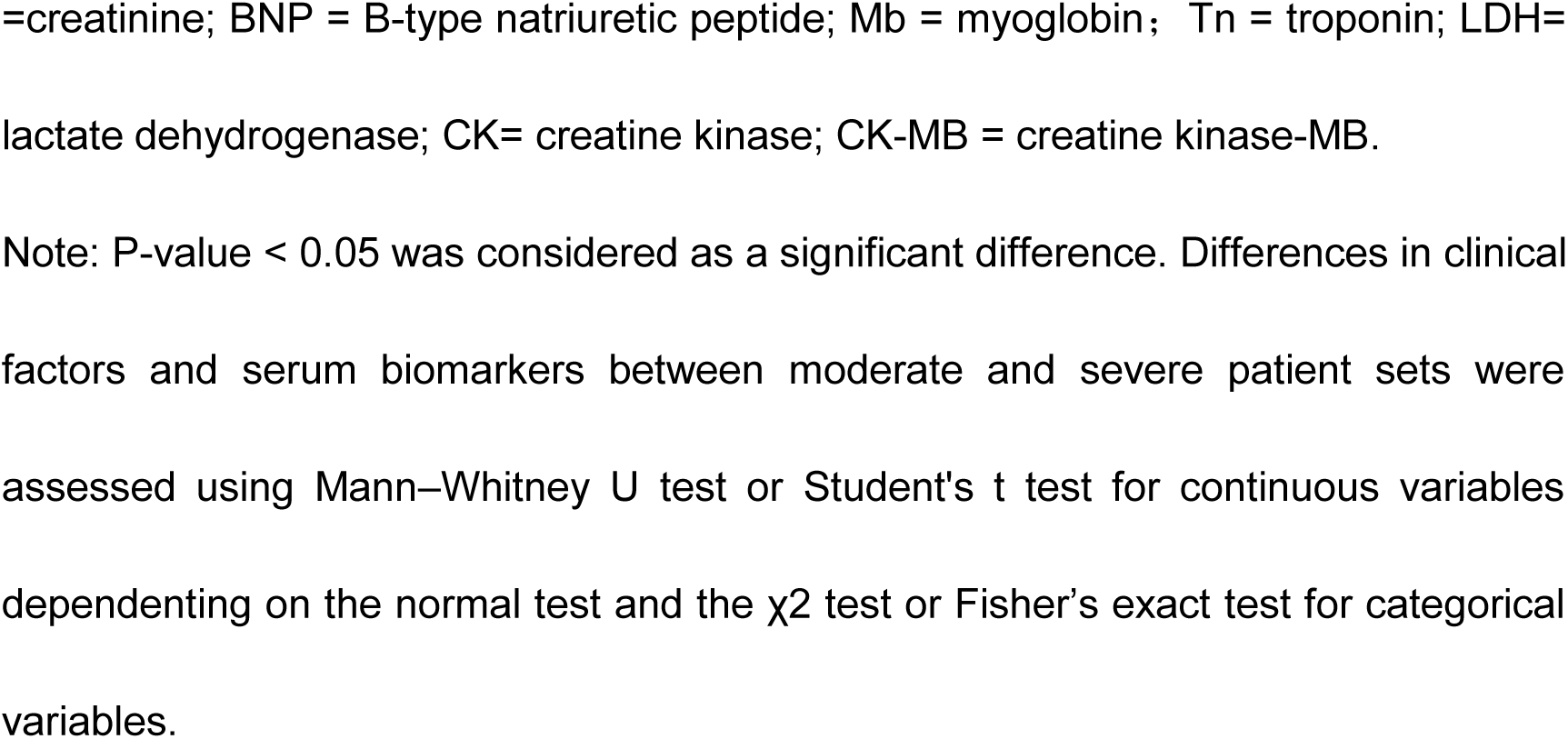
Clinical characteristics of patients in training and validation set (n=1209)

The auto-segmentation framework construction, radiomics feature extraction, feature selection, and predicting model building were established on the Research Portal V1.1 (Shanghai United Imaging Intelligence, Co., Ltd.).

### Adrenal gland and periadrenal fat auto-segmentation framework

For adrenal gland segmentation, we manually delineated bilateral adrenal glands from the CT images of 315 patients, 265 of them were used for training. The remaining data from 50 patients were used to evaluate the performance and the segmentation model yielded average Dice values of 79.48% for the left adrenal gland and 78.55% for the right adrenal gland. The entire adrenal gland achieved an average Dice value of 79.02%. Representative auto**-**segmentation results are shown in Figure 1. We also visually verified the segmentation results, and they were considered satisfactory based on radiologists’ judgment. The segmentation algorithm was then used to segment all the remaining data automatically.

**Figure 1.**
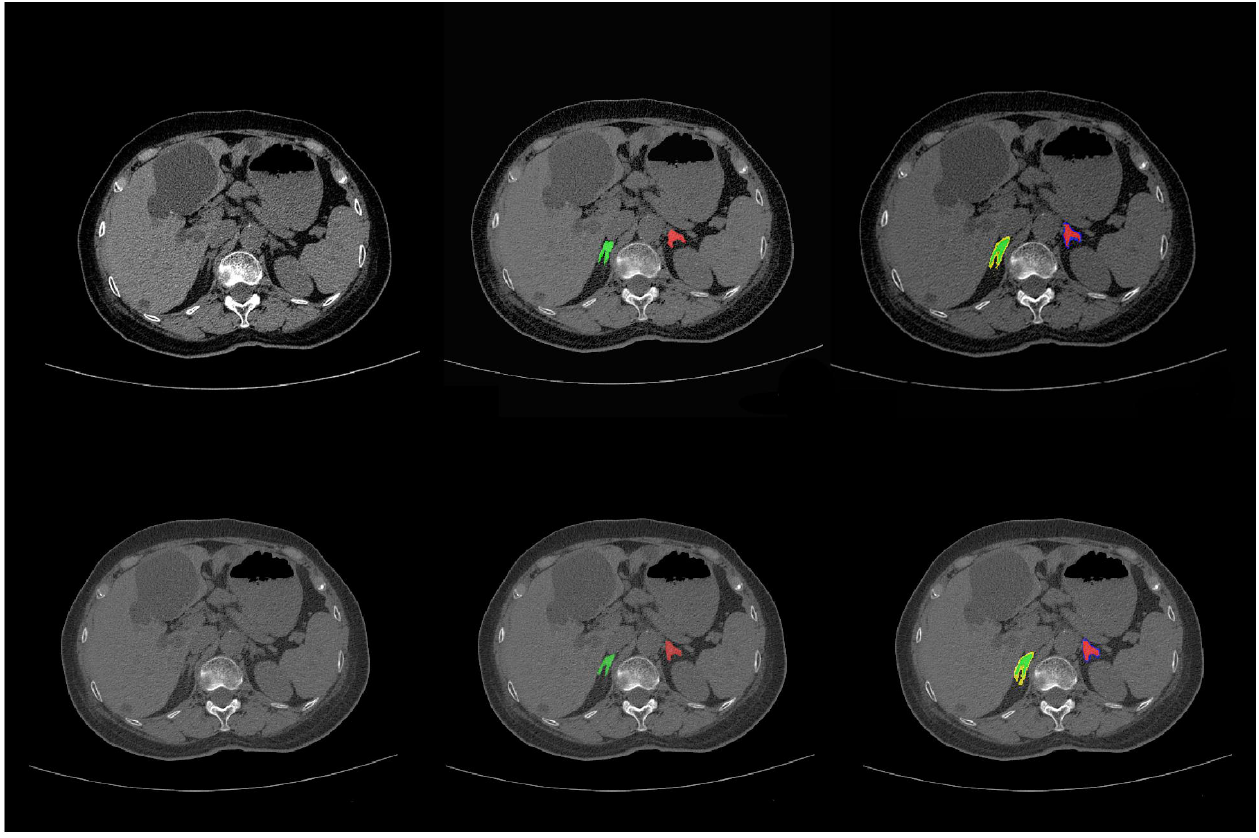
The comparison of auto-segmentation results and ground truth on representative cases from training set. The upper line shows the ground truth, lower line the auto-segmentation results. Green shows right adrenal glands and red shows left. Yellow shows right periadrenal fat and purple shows left.

### Radiomics feature and clinical indicator selection

In the training set, K-best and least absolute shrinkage and selection operator (LASSO) analyses were used to select the radiomics features most relevant to the progression of COVID-19. The number of radiomics features was reduced to 23 for building AM that included 8 first-order features and 15 texture features (Gray Level Co-occurrence Matrix [GLCM] = 3, Gray Level Size Zone Matrix [GLSZM] = 8, Gray Level Run Length Matrix [GLRLM] = 2 and Gray Level Dependence Matrix [GLDM] = 2); 68 for PM that included 11 first-order features, 2 sharp feature, and 55 texture features (GLCM = 13, GLSZM = 25, GLRLM = 7, GLDM = 9 and Neighboring Gray Tone Difference Matrix [NGTDM] = 1) and 82 for FM that included 12 first-order features and 70 texture features (GLCM=17, GLSZM=38, GLRLM = 3, GLDM=5 and NGTDM = 7). These features were evaluated to construct three radiomics models.

A total of 30 clinical factors and serum biomarkers were analyzed in our study. They were age, sex, IL-6, WBC, L, N, hemoglobin (HB), red blood cell count (RBC), blood platelet count (PLT), CRP, PCT, PT, thrombin time (TT), fibrinogen (FIB), DD, BS, Alanine transaminase (ALT), AST, total protein (TP), ALB, total bilirubin (TBil), D-Bil, blood urea nitrogen (BUN), creatinine (Cre), BNP, myoglobin (Mb), troponin (Tn), LDH, creatine kinase (CK), and CK-MB (Table 1). Next, 17 clinical factors and serum biomarkers were selected using univariate logistic regression analysis, and 7 indicators, LDH, L, HB, DD, WBC, TT, and TP, were selected using multivariate logistic regression analysis. The relationship between RadScore from FM used in the construction of radiomics nomogram (RN) and 30 clinical factors plus serum biomarkers were analyzed using Pearson correlation between training, validation, and test sets (Figure 2). The difference in RadScores with clinical factors or serum biomarkers was not significant. The radiomics information extracted from onset CT images belonged to another dimension, and this information was not affected by clinical factors and serum biomarkers. Then, 17 clinical factors and serum biomarkers were selected using univariate logistic regression analysis, and 7 indicators, LDH, L, HB, DD, WBC, TT, and TP, were selected using multivariate logistic regression analysis.

**Figure 2.**
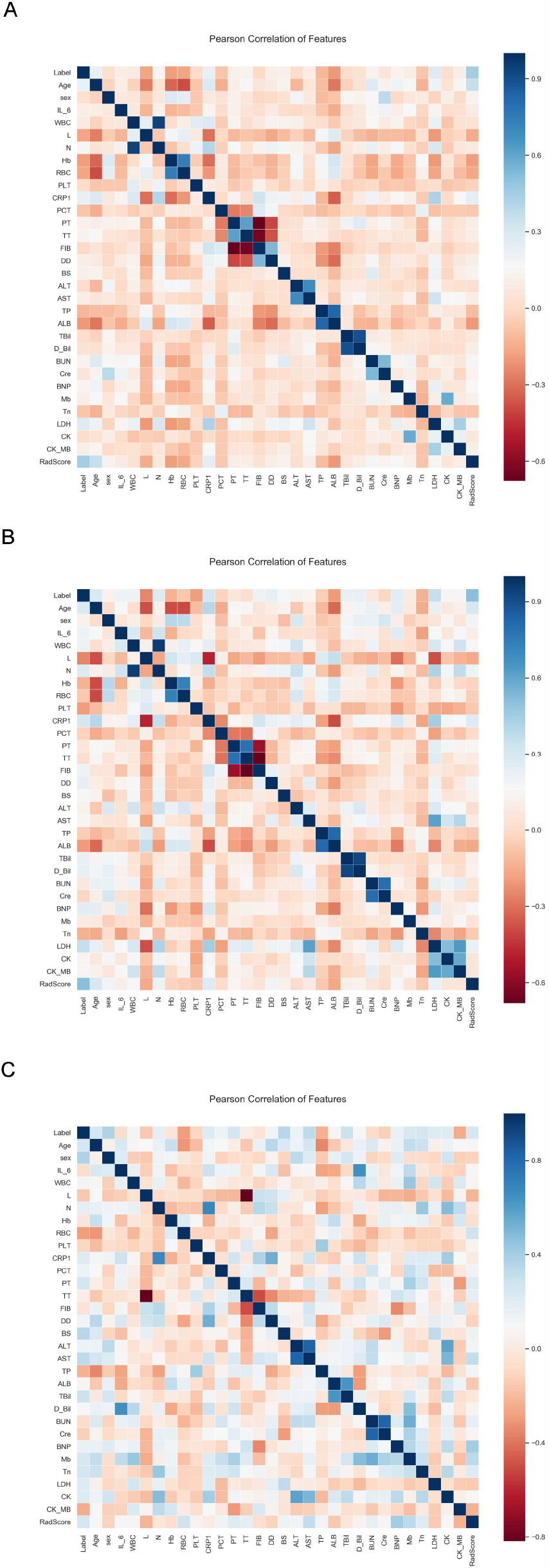
Heatmap in training, validation and test sets. FM = fusion of adrenal gland and periadrenal fat model; CM = clinical model; RN = radiomics nomogram.

### Three radiomics models and clinical model building

We developed three radiomics models (adrenal gland model [AM], periadrenal fat model [PM], a fusion of adrenal gland and periadrenal fat model [FM]) based on radiomics features and a clinical model (CM) based on the seven selected independent predictive clinical indicators. We used three evaluation indicators (area under the curve [AUC], 95% CI, sensitivity [SEN] and specificity [SPE]) to assess AM, PM, FM, and CM for predicting progression of patients with COVID-19 in training, validation, and test sets. In general, AM achieved an AUC of 0.692, 0.716 and 0.659 in the training set, validation set and test set respectively; PM achieved an AUC of 0.763, 0.736 and 0.645; FM achieved an AUC of 0.791, 0.760 and 0.686; CM obtained an AUC of 0.712, 0.717 and 0.692(Figure 3, Table 2).

**Table 2.** Predictive performances of models in training, validation and test set.

| Variable | Training set |  |  | Validation set |  |  | Test set |  |  |
| --- | --- | --- | --- | --- | --- | --- | --- | --- | --- |
|  | AUC(95%CI) | SEN | SPE | AUC(95%CI) | SEN | SPE | AUC(95%CI) | SEN | SPE |
| AM | 0.692(0.662-0.721) | 0.643 | 0.652 | 0.716(0.655-0.772) | 0.638 | 0.656 | 0.659(0.482-0.808) | 0.696 | 0.539 |
| PM | 0.763(0.735-0.789) | 0.651 | 0.749 | 0.736(0.675-0.790) | 0.654 | 0.664 | 0.645(0.469-0.797) | 0.652 | 0.692 |
| FM | 0.791(0.763-0.816) | 0.672 | 0.740 | 0.760(0.701-0.813) | 0.682 | 0.681 | 0.686(0.510-0.830) | 0.696 | 0.769 |
| CM | 0.712(0.682-0.740) | 0.613 | 0.700 | 0.717(0.655-0.772) | 0.676 | 0.724 | 0.692(0.517-0.835) | 0.872 | 0.615 |
| RN | 0.806(0.780-0.831) | 0.657 | 0.775 | 0.833(0.780-0.878) | 0.722 | 0.791 | 0.773(0.603-0.895) | 0.826 | 0.846 |

ACC = accuracy; SEN = sensitivity; SPE = specificity; AM = adrenal gland model; PM = periadrenal fat model; FM = fusion of adrenal gland and periadrenal fat model; CM = clinical model; RN = radiomics nomogram

**Figure 3.**
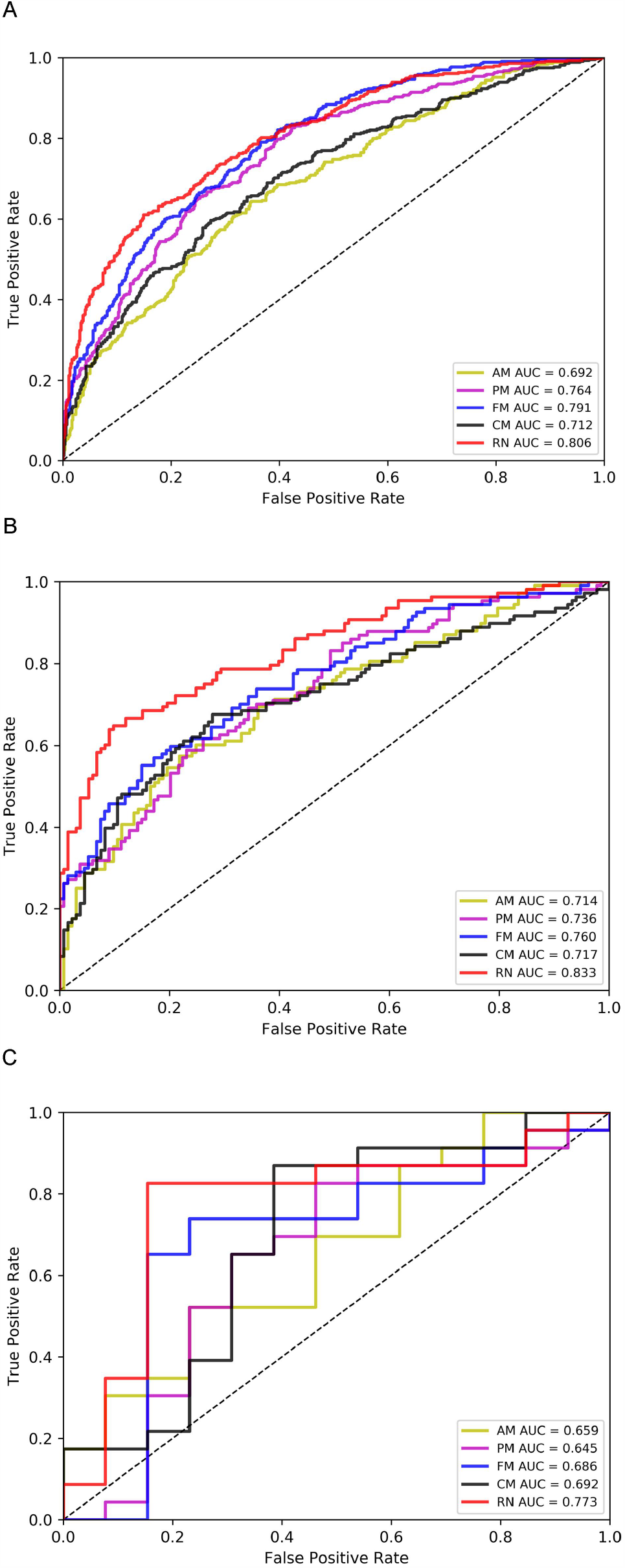
ROC curves in training, validation and test sets. AM = adrenal gland model; PM = periadrenal fat model; FM = fusion of adrenal gland and periadrenal fat model; CM = clinical model; RN = radiomics nomogram.

Box plots summarizing the RadScores and coefficients of seven clinical indicators in training, validation, and test sets directly demonstrate the difference between RadScore and coefficients of seven clinical indicators between the moderate and severe patient sets (Figure 4).

**Figure 4.**
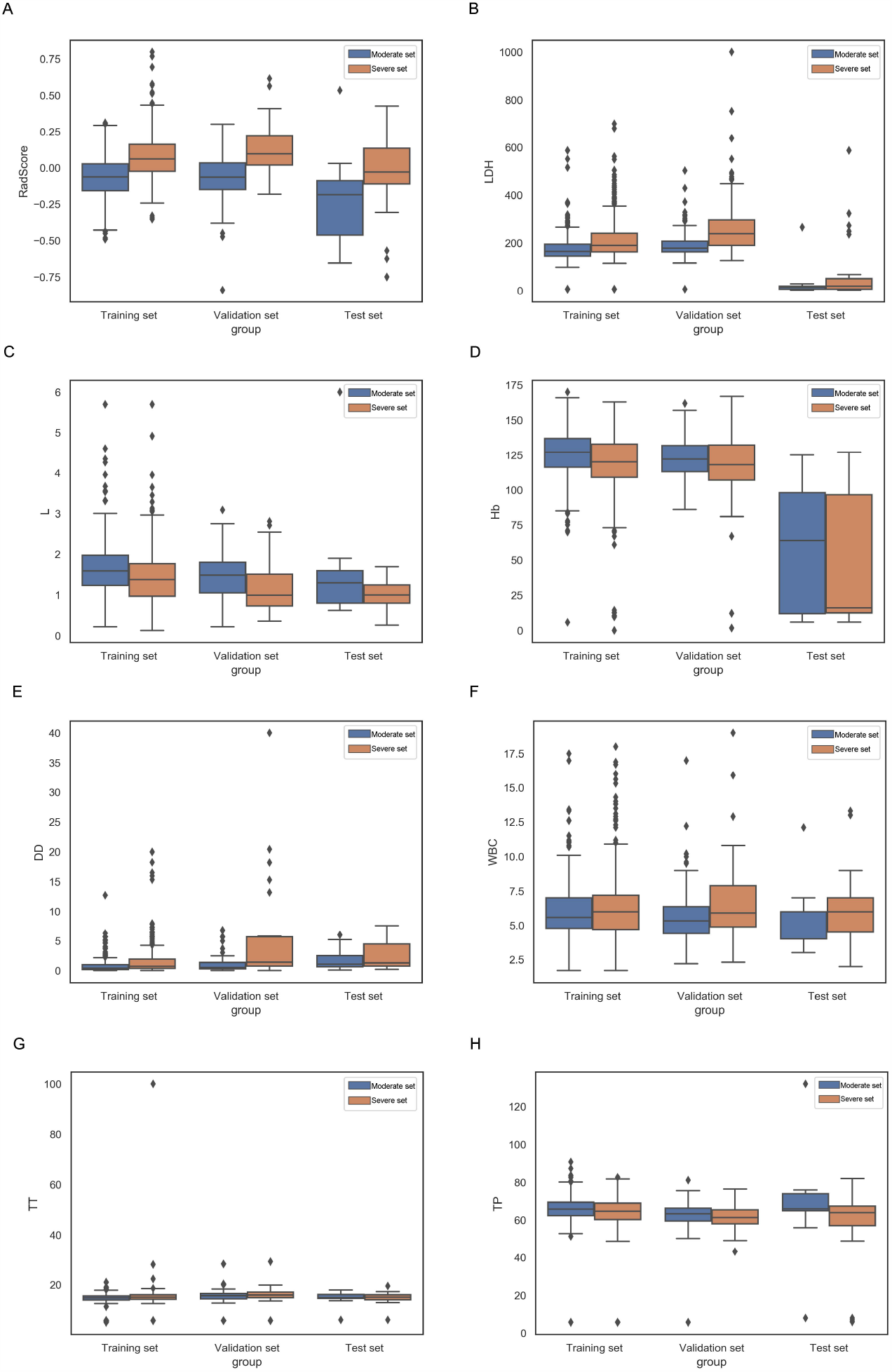
Box plots summarizing radscore and 7 clinical indicators between moderate patient and severe patient sets in training, validation and test sets. In each box plot, the horizontal line crossing the box is the median, the bottom and top of the box are the lowerand upper quartiles. FM = fusion of adrenal gland and periadrenal fat model; CM = clinical model; RN = radiomics nomogram; LDH = lactate dehydrogenase; L = lymphocyte count; Hb = hemoglobin, DD = D dimer; WBC = white blood cells count; TT = thrombin time and TP = total protein.

### RN construction and validation

Multivariate analysis revealed that RadScore and seven clinical indicators were significant independent factors predicting disease progression in patients with COVID-19. Using collinearity diagnosis, variance inflation factor (VIF) for the radiomics score and seven clinical indicators were from 1.007 to 1.191, indicating no severe collinearity in these factors. Next, we used the RadScore from FM combined with seven clinical indicators to construct the RN to assess disease progression in patients with COVID-19 (Figure 5). The RN showed satisfactory performance for predicting and assessing progression in patients with COVID-19 with AUC of 0.806 (95% CI, 0.780 to 0.831) in the training set, 0.833 (95% CI, 0.780 to 0.878) in the validation set, and 0.773 (95% CI, 0.603 to 0.895) in the test set (Figure 3, Table 2).

**Figure 5.**
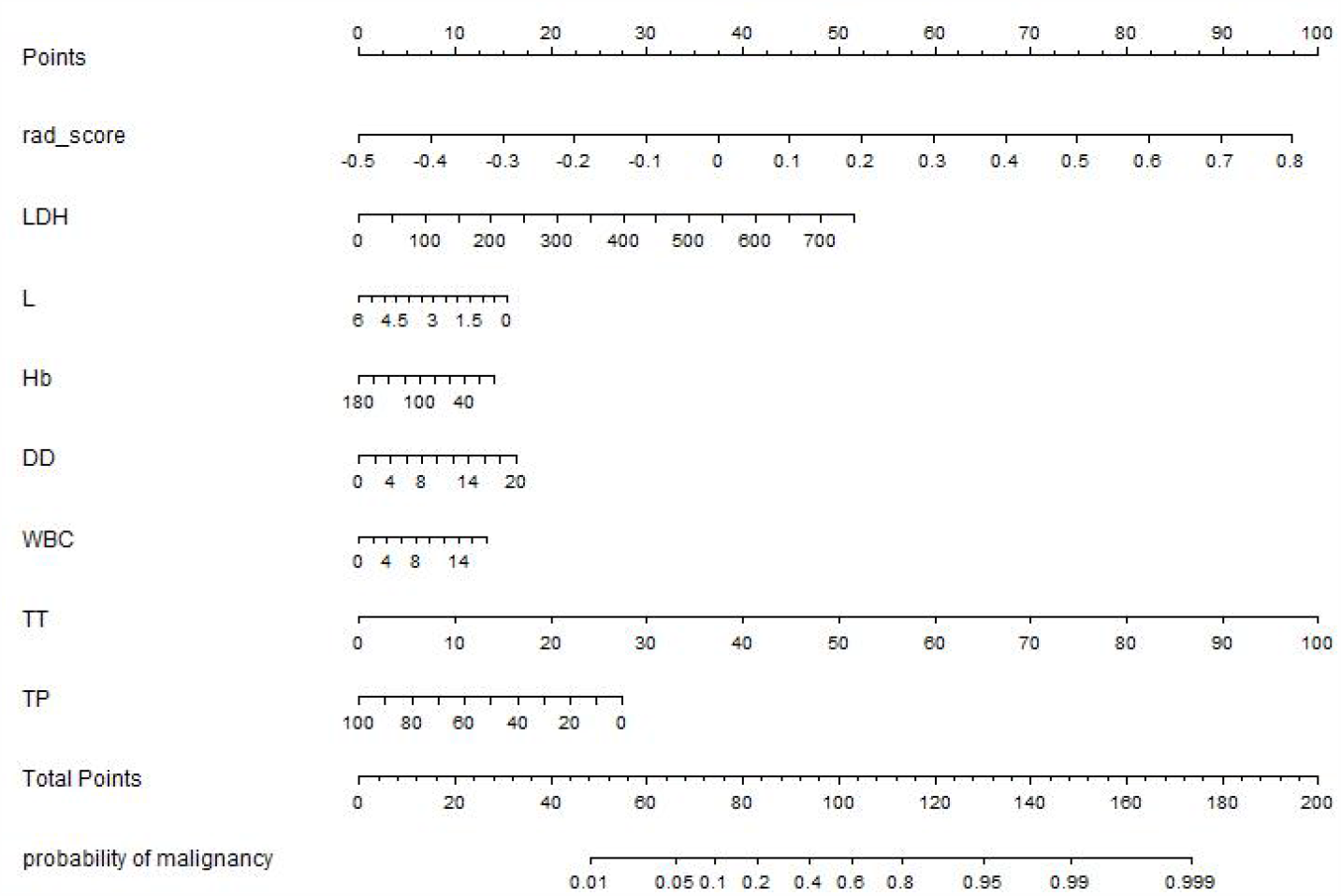
Radiomics nomogram developed in training set with radiomics features, lactate dehydrogenase(LDH), lymphocyte count(L), hemoglobin(Hb), D dimer(DD), white blood cells count(WBC), thrombin time(TT) and total protein(TP). Points are assigned for each variable by drawing a line upward from the corresponding variable to the Points line. The sum of points plotted on the total Points line corresponds with the severity of patients with COVID-19.

DeLong’s test was used to compare the AUCs of three radiomics models, CM and RN, in the training set. The result showed that the RN and FM were significantly better than CM (*P* < 0.0001). The difference between FM and RN was not statistically significant (*P* = 0.233) in the validation and test sets.

The calibration curve showed an agreement between the predicted and actual values. The Hosmer–Lemeshow test was not significant in the validation set (mean absolute error [MAE] = 0.075) or test set (MAE = 0.04), which suggests that there was no significant departure from actual values (Figure 6). Decision curve analysis (DCA) was used to evaluate the performance of RN (Figure 7). If the threshold probability was more than 0.3 in the validation set, the RN could get more net benefits than FM and CM. If the threshold probability was between 0.4 and 0.8 in the test set, RN can still get more net benefits than FM and CM.

**Figure 6.**
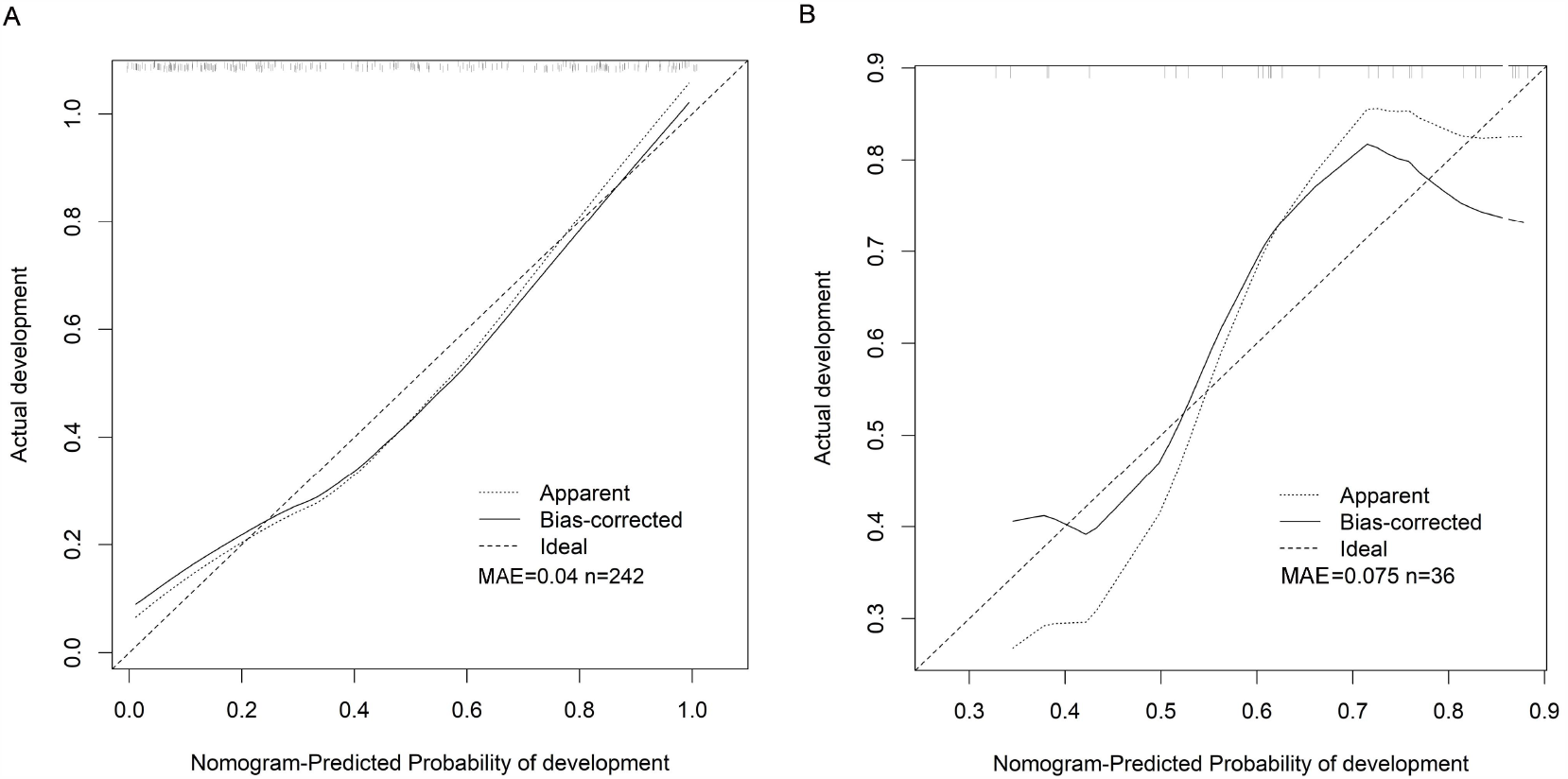
Calibration curves of the radiomic nomogram for predicting the disease progression of COVID-19 pneumonia in validation set (a) and test set (b). The y-axis represents the actual probability of the COVID-19 pneumonia becoming severe, the x-axis represents the predicted risk. Dashed line was reference line where an ideal nomogram would lie. Dotted line was the performance of radiomics nomogram, while the solid line corrects for any bias in radiomics nomogram. MAE is 0.04 in validation set and 0.075 in test set.

**Figure 7.**
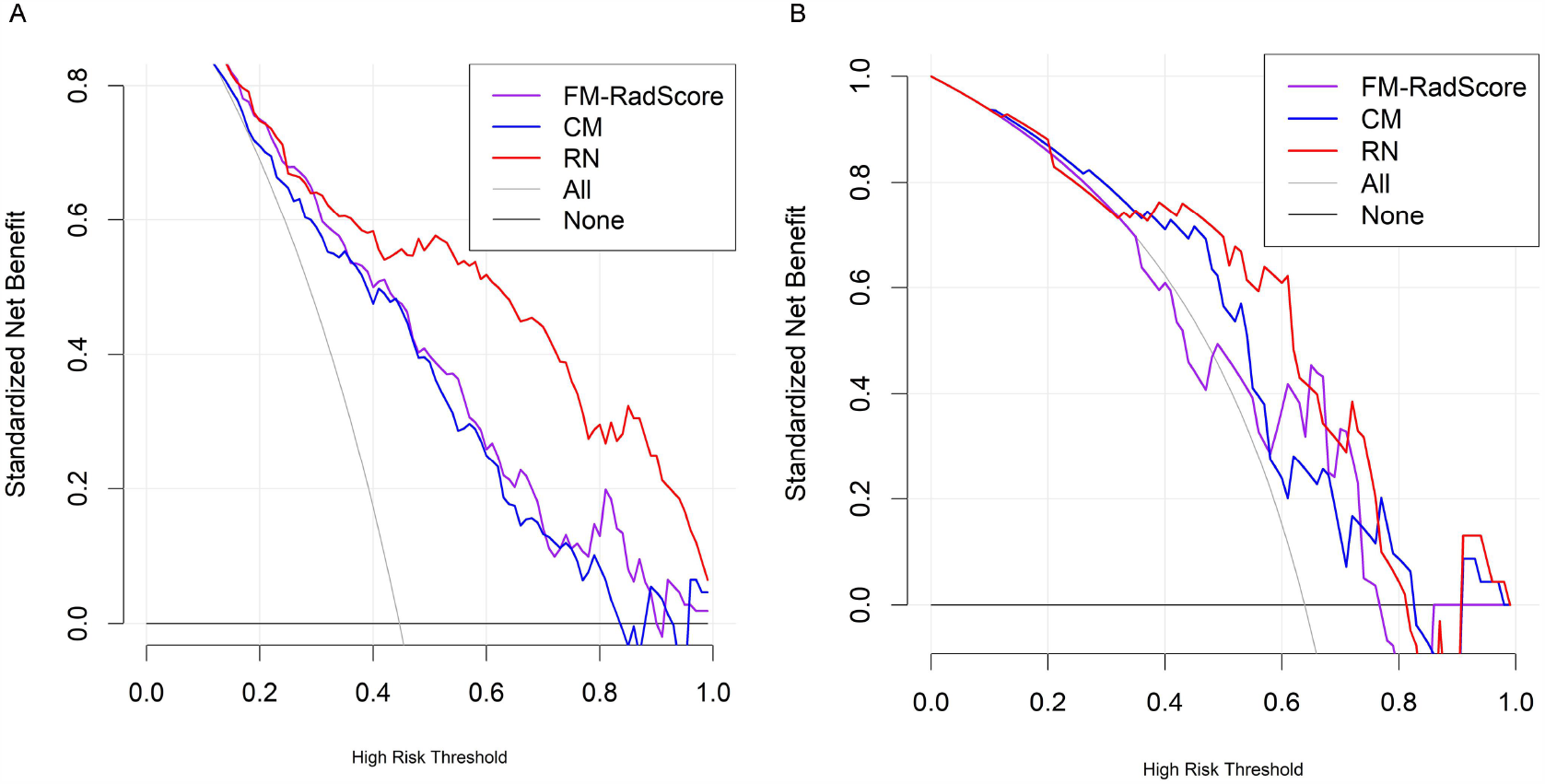
Decision curve analysis(DCA) for the radiomic model, clinical model and radiomics nomogram. The y-axis measures the net benefit. using the clinical model,radiomic model and radiomics nomogram in the study to predict COVID-19 pneumonia progress adds more benefit than the treat all patients as severity patients scheme or the treat none scheme. The net benefit of radiomics nomogram was better than clinical model and radiomic model in both two sets and with several overlaps in the training set.

## Discussion

In this study, an adrenal gland auto-segmentation framework was constructed. We rapidly finished the auto-segmentation of adrenal glands and periadrenal fat using the framework. We found several radiomics features and seven clinical indicators related to disease progression in patients with COVID-19. Next, we built and validated a RN for predicting disease progression based on radiomics features extracted from the onset of the adrenal gland and periadrenal fat CT images combined with clinical indicators. Our study indicated that the auto-segmentation framework could robustly localize the adrenal glands and accurately refine their boundary. The RN using adrenal glands and periadrenal fat onset CT images performed well, reflecting that the microscopic changes in adrenal glands and periadrenal fat in patients with COVID-19 can be detected using radiomics features.

Our results suggested that adrenal gland and periadrenal fat changes on the onset of CT images in severe patients differed from those in moderate patients. Results from autopsies in 10 patients from COVID-19 performed by Zinserling et al. (14) showed that even macroscopic changes of adrenal glands were unremarkable. However, they found inflammation—small proliferations of cells with enlarged light nuclei and mononuclear infiltration, CD3+ and CD8+ in different layers of adrenal glands and their surrounding tissue, such as periadrenal fat. These changes may be related to direct damage of adrenal glands and indirect changes stimulated by systemic inflammation caused by SARS-CoV-2 or immune state. In one respect, the changes were caused by direct damage done by SARS-CoV-2. The SARS virus has been identified in adrenal cells, suggesting a direct local replication-mediated cytopathic effect of the virus in adrenal tissue (15). Moreover, virus may cause hemorrhage, necrosis, or thrombosis at the adrenal level. (16) Adrenal lesions due to influenza and other viruses have been described previously (17). Recent findings have indicated the possibility of venous thrombo-embolism in patients with COVID-19 that may cause an acute adrenal insufficiency that may be an indicator of making the disease worse (18). Additionally, the adrenal gland is one of the most highly vascularized organs of the human body, with almost every adrenal cell in close location to endothelial cells. Therefore, endotheliitis in COVID-19 may further increase the vulnerability of adrenal tissue. However, this needs further study and validation. We hypothesized that the same effect could occur in patients with COVID-19 due to similar virus types. Our predicting results showed that AM’s AUC value was slightly better than PM’s, but there was no statistical significance. That may indicate that the inner changes in adrenal glands were not different from the changes from periadrenal fat. Further research is needed to confirm changes in periadrenal fat were caused by adrenal gland inflammatory cells’ infiltration or direct damage of SARS-CoV-2 (19).

Previous research also reported that rather than direct damage by SARS-CoV-2 itself, adrenal gland changes were believed to be caused by immune overreaction or cytokine storm-inducing endocrinological pathway impaired through a tight binding mechanism of SARS-CoV-2 and ACE2 (20). Endocrinological pathway impairment may be related to the following aspects. First, ACE2 is highly expressed in human adrenals, which serves as the entry receptor for SARS-CoV-2 that is bound to be significantly affected. Although no studies have proved it, some work has proposed that the imbalanced ACE/ACE2 axis may mediate tissue repair and wound healing pathways in the lungs, such as fibrosis after SARS-CoV-2 (21). Second, the main substrate for ACE2 is angiotensin II. ACE2 acts as a negative regulator of the renin-angiotensin-aldosterone system (RAAS) by converting the active angiotensin and angiotensin II to the inactive angiotensin 1-7 (22). The adrenal gland is one of the most critical end organs of RAAS that is fundamental in regulating blood pressure, maintaining homeostasis, and is mediated in the organs’ inflammatory response. Finally, the SARS virus contains several permutations of amino acid sequences with homology to the antigenic relevant residues of ACTH (23) that will have potential and significant pathophysiological effects due to molecular mimicry. ACTH is a key hormone that regulates autoantibodies’ release by adrenal corticosteroids and could abrogate the adrenal stress response when autoantibodies were binding ACTH. Potential cross-reactivity of antibodies may cause local infiltration with immunocompetent cells in adrenal glands(14). In summary, these aspects suggested that adrenal cells and surrounding tissue damage in patients with COVID-19 may be caused by a viral infection and further secondary inflammatory plus autoimmune processes located in the adrenal glands.

In conclusion, we proposed and constructed an adrenal gland auto-segmentation framework based on chest CT images and AI technology. We automatically obtained the ROI from the onset CT images through the auto-segmentation framework and inflation algorithm, extracted the radiomics features, and then developed and validated a RN for predicting the disease progression of COVID-19 combining onset CT images with clinical indicators. Until now, we did not find any research using adrenal gland CT parameters as indicators to evaluate the progression in patients with COVID-19, especially in the radiomics field. Our work has some limitations. First, we used the chest CT images as data resources. However, taking clinical practicality and radiation to patients into consideration, there is no need to perform another CT scan using professional adrenal glands CT parameters to observe adrenal lesions a little better because CT examination of patients with COVID-19 is mainly to detect and observe pulmonary lesions. Second, different from other studies, we selected the entire organ as ROIs rather than the lesion itself. The periadrenal fat area may not be precise and contain some other tissue that is indistinguishable to the human eye. Although there are some deficiencies, our findings validated the potential for radiomics features extraction from adrenal glands and periadrenal fat CT images to be the indicators of COVID-19 prognosis. However, these need to be validated in large-scale prospective studies.

## Methods

### Data resources and image grouping

CT images and clinical data were retrospectively and consecutively collected from two hospitals: Huoshenshan Hospital (HSH) (n = 1,209) and Maternal and Child Health Hospital Optical Valley Branch Hospital of Hubei province (MCH) (n = 36), China. Diagnosis and clinical classification of patients with COVID-19 were confirmed according to Diagnosis and Treatment Protocol for Novel Coronavirus Pneumonia (Trial Version 8). Patients of age ≤ 14 years (n = 2) were excluded. All images were non-enhanced chest CT images and reconstructed at a slice thickness of 1.00 mm. Details of CT characteristics are listed in Supplementary Table. Patients with adrenal lesions were excluded after being evaluated by two radiologists with more than 10 years of experience. We chose chest CT images scanned within four days to the first diagnosis as the onset image. If the CT scan was done more than once, we chose the one closer to the admission date. Figure 8 demonstrates the inclusion and exclusion criteria. Figure 9 shows the workflow of our study.

**Figure 8.**
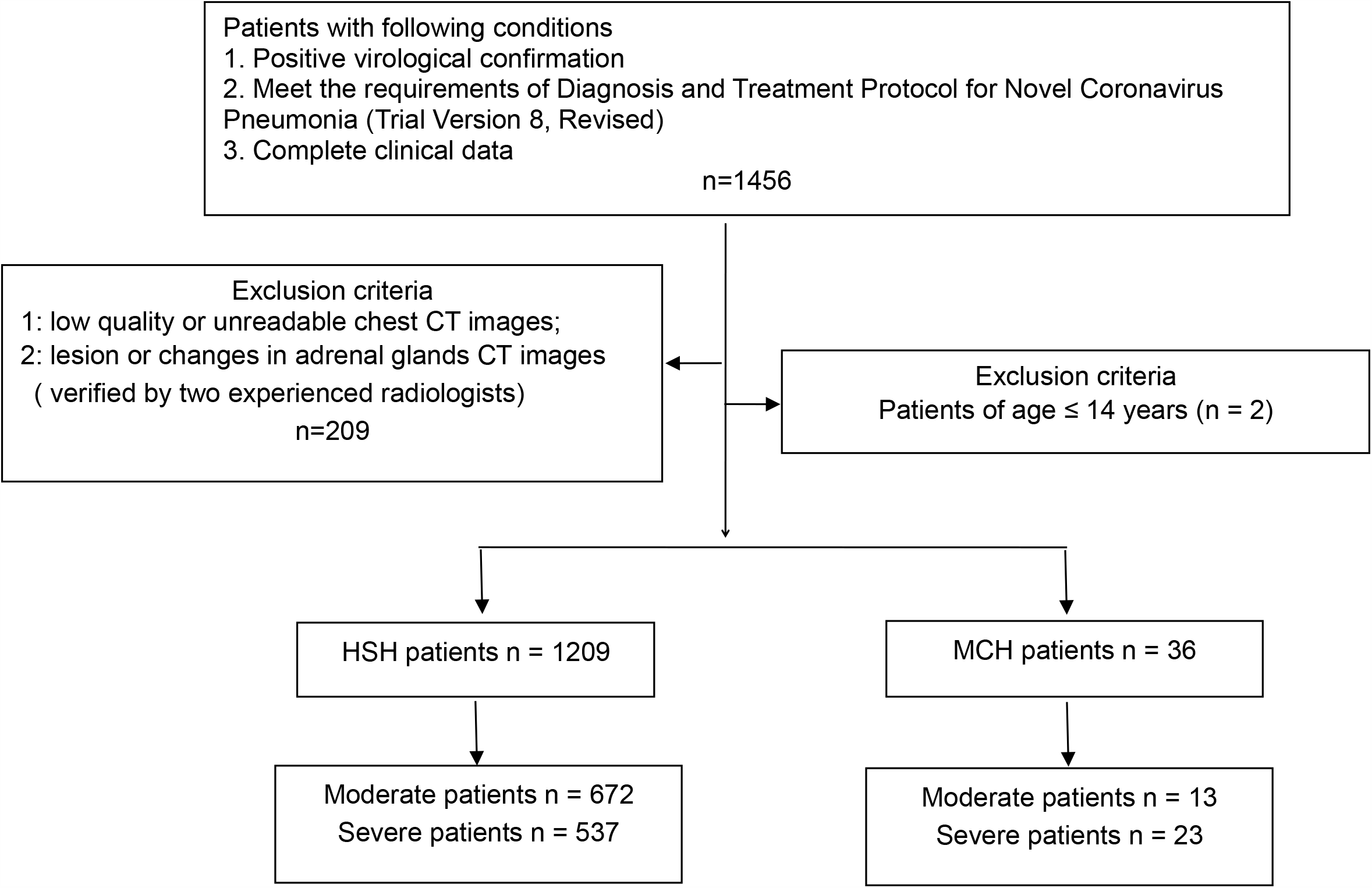
Flowchart shows the patient inclusion and exclusion criteria

**Figure 9.**
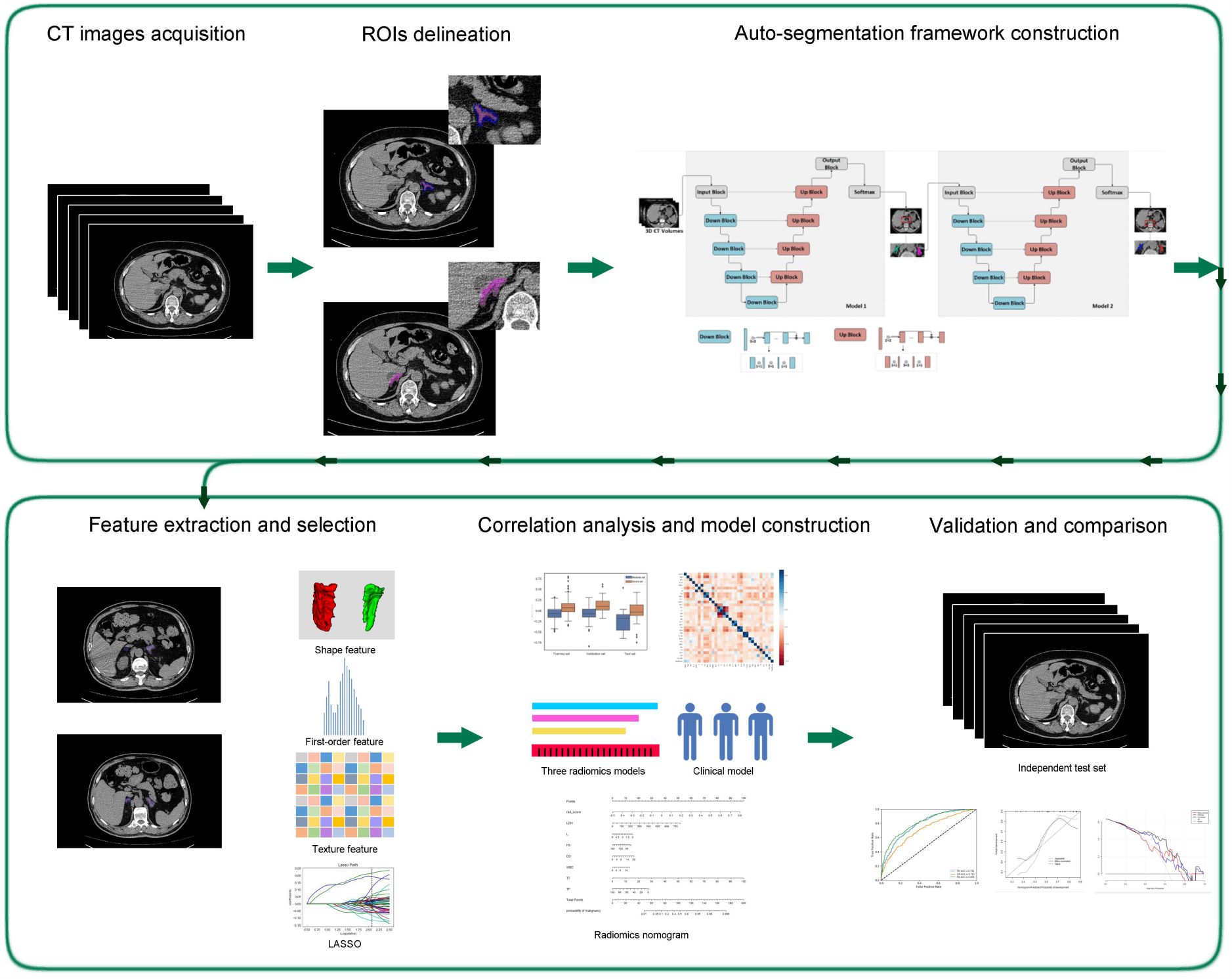
The workflow of study

CT images of 1,209 patients from Huoshenshan hospital were used to construct the prediction models divided into the training set (967/1209, 80%) and the validation set (242/1209, 20%) randomly. We used different imaging and clinical features and their combinations to predict the progression of COVID-19. We constructed five models: adrenal gland radiomics model (AM), periadrenal fat radiomics model (PM), a fusion of the adrenal gland and periadrenal fat radiomics model (FM), clinical model (CM) and radiomics nomogram (RN). A total of 36 Patients (13 moderate and 23 severe patients) from Maternal and Child Health Hospital Optical Valley Branch Hospital were included as an independent test set.

### Auto-segmentation framework and ROIs delineation

The objective of image segmentation is to extract ROIs, including bilateral adrenal glands and the periadrenal fat. Here, we propose a cascaded V-net network framework to segment the bilateral adrenal glands automatically, and ROI of periadrenal fat was obtained using the inflation algorithm based on the adrenal gland. An experienced radiologist (Y.F.) manually delineated the ROIs of bilateral adrenal glands which were as the ground truth labels of adrenal, and he was asked to delineate the adrenals according to the image delineating principles of BraTS 2018. For the auto-segmentation framework, we first trained a coarse localization model as Model 1, which can perform coarse segmentation to locate the adrenal gland area. The second V-Net model was used for defining segmentation as Model 2, and further divided into the left adrenal gland and the right adrenal gland.

The main structure of V-net includes Down-Block and Up-Block. The high-level context information was extracted by convolutions and Down-Blocks first. Then, skip connections were used to fuse high-level context information and fine-grained local information. Through residual function and skip connection, the model can achieve high accuracy. Normalization is performed before the image is input to the network. The segmentation frame is shown in Figure 10. The cascade model segmentation of the bilateral adrenal glands worked sequentially to obtain more accurate segmentation results.

**Figure 10.**
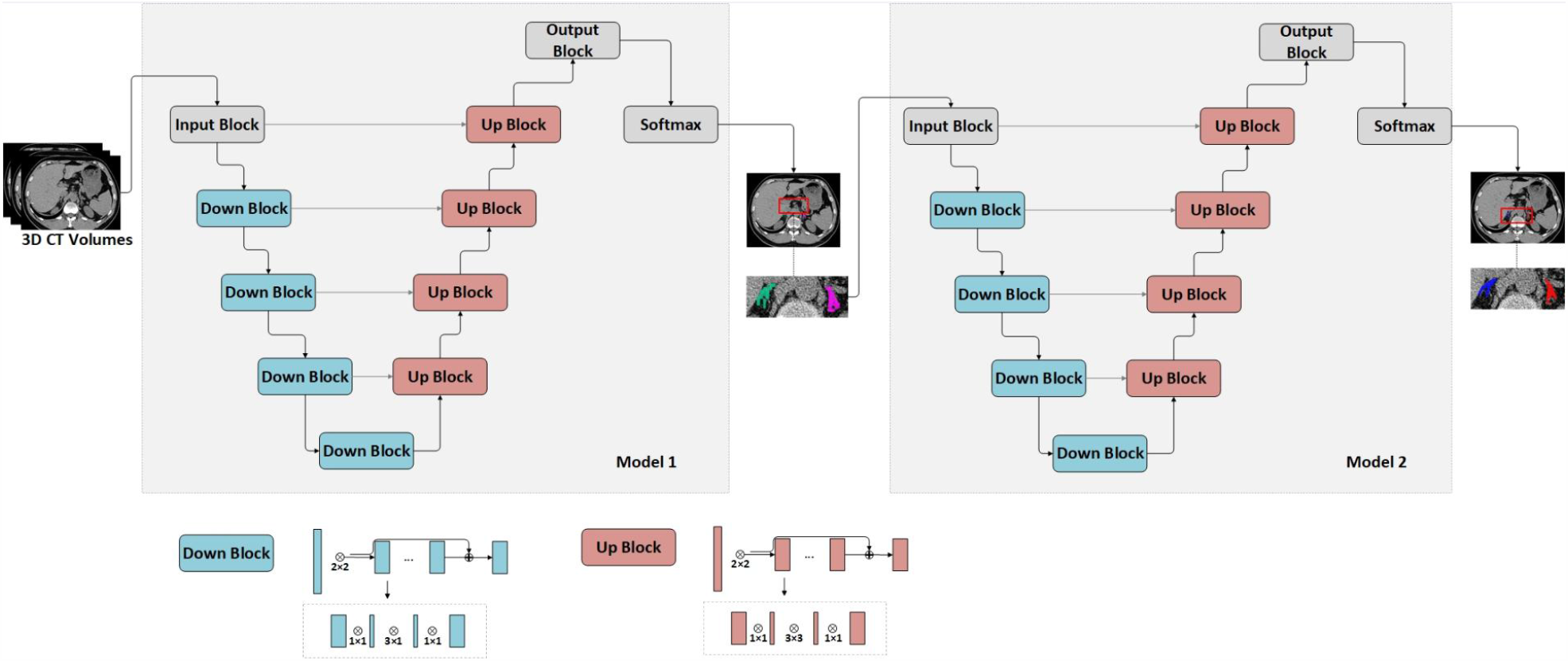
The framework of the used V-net

ROIs including bilateral adrenal glands and periadrenal fat from all CT images without annotation were segmented using the auto-segmentation framework. Next, the ROIs with serious nonconformity were manually verified by two radiologists with more than 5 years of clinical experience and validated by another radiologist with 20 years of clinical experience.

### Radiomics feature selection and models building

Radiomic features were extracted from ROIs on the CT images using a Python package (PyRadiomics V3.0). B-spline interpolation resampling was used to normalize the voxel size, and the anisotropic voxels were resampled to form isotropic voxels of 1.0 mm × 1.0 mm × 1.0 mm in the feature extraction. A total of 2,264 features of each ROI were extracted, which can be divided into 3 common feature groups, 450 first-order features, 14 shape features, and 1,800 texture features (gray-level co-occurrence matrix [GLCM] = 525, gray-level run-length matrix [GLRLM] = 350, gray-level size zone matrix [GLSZM] = 400, gray-level dependence matrix [GLDM] = 400 and neighboring Gray Tone Difference Matrix [NGTDM] = 125) (24). Next, the feature in the training set was preprocessed by standardization. The mean and variance in the training set were applied to the validation and test sets.

To reduce dimensionality and select significant features, we performed a feature dimension reduction process to select the most relevant features. First, a univariate analysis named K-best was employed (25). It selected features according to K highest scores computed through ANOVA F-value and P-value between label and features. Features with a significant difference (*P* < 0.05) were selected. Next, the LASSO feature-selection algorithm was used to extract the most informative radiomics features to prevent the “curse of dimensionality”. After feature extraction and selection, logistic regression (LR) algorithms were trained to construct three radiomics models (AM, PM, and FM) for predicting the disease progression of COVID-19 using a five-fold cross-validation strategy. In the FM model, the RadScore of the patient was calculated according to the LASSO algorithm.

### RN construction and evaluation

We used univariate analysis to assess the relationship between clinical factors plus serum biomarkers and disease outcome. Features with *P* < 0.05 were introduced into multivariate LR analysis with radiomics features to select the best combination using a five-fold cross-validation strategy. The best assignment of training and validation sets was chosen for the next analysis. Next, we applied the multivariate LR model to build CM using valuable clinical indicators and RN using the RadScore from FM with clinical indicators to predict the disease progression of COVID-19.

We conducted collinearity diagnosis by calculating the VIF for variables in RN to detect multicollinearity among the radiomics nomogram variables. In the end, RN was verified in the validation and test sets. Calibration curves and Hosmer–Lemeshow test were used to assess the relation between the predicted risks and actual results. DCA was used to evaluate the performance of the RN.

### Statistics

Before model building, differences in clinical factors and serum biomarkers between moderate and severe patient sets were assessed using Mann-Whitney U test or Student’s *t*-test for continuous variables and the χ2 test or Fisher’s exact test for categorical variables. We analyzed all data using SPSS for Windows version 26.0 (IBM Corp., Armonk, New York, USA). *P* < 0.05 was considered a statistically significant difference.

Dice value was used to assess the effectiveness of auto-segmentation framework. The AUC of receiver-operating characteristics (ROC) with 95% confidence interval (95% CI), sensitivity, and specificity were used to evaluate the performance of AM, PM, FM, CM and RN. Accuracy was calculated to assess the prediction performance. Differences in AUC values among different models were estimated using the DeLong test.

## Data Availability

The database used and/or analysed during the current study are available from the corresponding author on reasonable request.

## Author contributions

First author, Mudan Zhang: design of the work, conducting experiments, analyzing data and writing the manuscript; Co-first author, Xuntao Yin: acquiring data, design of the work, supervising the work and review the manuscript.

Yan Zha: design of the work; Xianchun Zeng: Resources; Wuchao Li: statistics analysis; Xiaoyong Zhang: Technology analysis; Jingjing Cui: statistics analysis; Jie Tian: Review & Editing.

All authors have approved the submitted version (and any substantially modified version that involves the author’s contribution to the study). All authors have agreed both to be personally accountable for the author’s own contributions and to ensure that questions related to the accuracy or integrity of any part of the work.

## Study approval

This multicenter study was approved by the ethics committees of Guizhou Provincial People’s Hospital (2020, NO.01). Because of its retrospective nature, the need to obtain informed consent from patients was waived. The study was performed according to the principles of the declaration of Helsinki.

## Acknowledgements

This study was supported Science and Technology Foundation of Guizhou Province (QKHZC[2020]4Y002,QKHPTRC[2019]5803), the Guiyang Science and Technology Project (ZKXM[2020]4), Guizhou Science and Technology Department Key Lab. Project (QKF[2017]25), Beijing Medical and Health Foundation (YWJKJJHKYJJ-B20261CS) and Special Fund for basic Research Operating Expenses of public welfare research institutes at the central level of Chinese Academy of Medical Sciences (2019PT320003).

---

**Supplementary table.** CT characteristics.

| Manufacturer | Scanner type | Ratation<br>time | Resolutio<br>n | tube<br>voltage<br>(kv) | tube<br>current<br>(mAs) | Section<br>thickness | Reconstructi<br>on technique | Algorithm |
| --- | --- | --- | --- | --- | --- | --- | --- | --- |
| SIEMENS | SOMATOM<br>Perspective | 0.6s | 512x512 | 110 | 49 | 1 | ADMIRE 3 | I50s |
| SIEMENS | SOMATOM<br>Definition | 0.5s | 512x512 | 100 | 100 | 1 | FBP | B50f medium sharp |
| SIEMENS | SOMATOM<br>Definition AS+ | 0.5s | 512x512 | 120 | 52 | 0.6 | FBP | B50f medium sharp |
| SIEMENS | SOMATOM<br>Force | 0.5s | 512x512 | 100 | 52 | 1 | ADMIRE 3 | BI57 |
| GE MEDICAL<br>SYSTEMS | BrightSpeed | 0.5s | 512x512 | 100-120 | 120-140 | 1.25 | FBP | chest,lung,stnd |
| GE MEDICAL<br>SYSTEMS | LightSpeed VCT | 0.5s | 512x512 | 100-120 | 120-140 | 0.625 | FBP | chest,lung,stnd |
| NMS | NeuViz 16<br>Essence | 0.5s | 512x512 | 120 | 100-120 | 1.25 | Iterate | lung |
| UIH | uCT 510 | 0.5s | 512x512 | 120 | 120-150 | 1.5 | FBP | KARL 3D®iterative<br>reconstruction |

## References

1. Tian M, Long Y, Hong Y, Zhang X, and Zha Y. The treatment and follow-up of‘recurrence’ with discharged COVID-19 patients: data from Guizhou, China. Environmental Microbiology. 2020;22(8):3588–92.

2. Giorgia Guglielmi. Fast coronavirus tests: what they can and can’t do. Nature. 2020; 585, 496–498.

3. Zhang K. Clinically Applicable AI System for Accurate Diagnosis, Quantitative Measurements and Prognosis of COVID-19 Pneumonia Using Computed Tomography. CELL. 2020; 181(6): 1423–1433.

4. Ting DSW, Carin L, Dzau V, and Wong TY. Digital technology and COVID-19. Nat Med. 2020. 26: 459–461

5. Agarwal S, Agarwal SK. Endocrine changes in SARS-CoV-2 patients and lessons from SARS-CoV. Postgrad Med J. 2020;96(1137):412–6.

6. Bellastella G, Maiorino MI, and Esposito K. Endocrine complications of COVID-19: what happens to the thyroid and adrenal glands? Journal of Endocrinological Investigation. 2020;43(8):1169–70.

7. Waldemar Kanczkowski, and Mariko Sue SRB, 2. The adrenal gland microenvironment in health, disease and during regeneration. 2017. 16(3):251–265.

8. Pal R, and Banerjee M. COVID-19 and the endocrine system: exploring the unexplored. Journal of Endocrinological Investigation. 2020;43(7):1027–31.

9. Peter Sever, Sebastian L Johnston. The Renin-Angiotensin system and SARS-CoV-2 infection: A role for the ACE2 receptor? [published online May 13, 2020]. J Renin Angiotensin Aldosterone Syst. doi: 10.1177/1470320320926911

10. Berger I, Werdermann M, Bornstein SR, and Steenblock C. The adrenal gland in stress – Adaptation on a cellular level. The Journal of Steroid Biochemistry and Molecular Biology. 2019;190:198–206.

11. Gillies RJ, Kinahan PE, Hricak H. Radiomics: Images Are More than Pictures, They Are Data. Radiology. 2016. 278(2):563–77.

12. Han, M., Yao, G., Zhang, W., Mu, G., Zhan, Y., Zhou, X., & Gao, Y. Segmentation of CT Thoracic Organs by Multi-resolution VB-nets. SegTHOR@ISBI. 2019. Corpus ID: 139100153

13. Hua R, Huo Q, Gao Y, Sui H, Zhang B, Sun Y, et al. Segmenting Brain Tumor Using Cascaded V-Nets in Multimodal MR Images. [published online Feb 14, 2020] Frontiers in Computational Neuroscience. doi: 10.3389/fncom.2020.00009.

14. Zinserling VA, Semenova NY, Markov AG, et al. Inflammatory Cell Infiltration of Adrenals in COVID-19. Horm Metab Res. 2020. 52(9):639–641.

15. Gu J, and Korteweg C. Pathology and Pathogenesis of Severe Acute Respiratory Syndrome. The American Journal of Pathology. 2007;170(4):1136–47.

16. Henry BM, Vikse J, Benoit S, et al. Hyperinflammation and derangement of renin-angiotensin-aldosterone system in COVID-19: A novel hypothesis for clinically suspected hypercoagulopathy and microvascular immunothrombosis. Clin Chim Acta. 2020;507:167–73.

17. Zinserling AV, Aksenov OA, Melnikova VF, Zinserling VA. Extrapulmonary lesions in influenza. Tohoku J Exp Med. 1983. 140(3):259–72.

18. Tseng YH, Yang RC, Lu TS. Two hits to the renin-angiotensin system may play a key role in severe COVID-19. Kaohsiung J Med Sci. 2020;36(6):389–92.

19. Battisti S, Pedone C, Napoli N, Russo E, Agnoletti V, Nigra SG, et al. Computed Tomography Highlights Increased Visceral Adiposity Associated With Critical Illness in COVID-19. Diabetes Care. 2020;43(10):e129–e30.

20. Datta PK, Liu F, Fischer T, et al. SARS-CoV-2 pandemic and research gaps: Understanding SARS-CoV-2 interaction with the ACE2 receptor and implications for therapy. Theranostics. 2020;10(16):7448–64.

21. Delpino MV, Quarleri J. SARS-CoV-2 Pathogenesis: Imbalance in the Renin-Angiotensin System Favors Lung Fibrosis. Front Cell Infect Microbiol. 2020;10:340.

22. Cheng H, Wang Y, and Wang GQ. Organ - protective effect of angiotensin - converting enzyme 2 and its effect on the prognosis of COVID-19. Journal of Medical Virology. 2020;92(7):726–30.

23. Pal R. COVID-19, hypothalamo-pituitary-adrenal axis and clinical implications. Endocrine. 2020;68(2):251–2.

